# Incidence of chikungunya virus infections among Kenyan children with neurological disease: a prospective cohort study

**DOI:** 10.1101/2021.05.27.20138404

**Authors:** Doris K. Nyamwaya, Mark Otiende, Lilian Mwango, Symon M. Kariuki, Berrick Otieno, Donwilliams O. Omuoyo, George Githinji, Barnes S. Kitsao, Henry K. Karanja, John N. Gitonga, Zaydah R. de Laurent, Alun Davies, Salim Mwarumba, Charles N. Agoti, Samuel M. Thumbi, Mainga M. Hamaluba, Charles R. Newton, Philip Bejon, George M. Warimwe

## Abstract

**Background:** Neurological complications due to chikungunya virus (CHIKV) infection have been described in different parts of the world, with children being disproportionately affected. However, the burden of CHIKV-associated neurological disease in Africa is currently unknown.

**Methods:** We estimated the incidence of CHIKV infection among children hospitalised with neurological disease in coastal Kenya. We used reverse-transcriptase polymerase chain reaction (RT-PCR) to systematically test for CHIKV in cerebrospinal fluid (CSF) samples from children aged <16 years hospitalised with symptoms of neurological disease at Kilifi County Hospital between January 2014 and December 2018. Clinical records were linked to the Kilifi Health and Demographic Surveillance System and population incidence rates of CHIKV infection estimated.

**Findings:** There were 18,341 paediatric admissions during the 5-year study period, of which 4,332 (24%) had CSF collected. The most common clinical reasons for CSF collection were impaired consciousness, seizures and coma (47%, 22% and 21% of all collections, respectively). After acute investigations done for immediate clinical care, CSF samples were available for 3,980 admissions, of which 367 (9.2%) were CHIKV RT-PCR positive. The annual incidence of CHIKV-associated neurological disease varied between 13 to 58 episodes per 100,000 person-years among all children <16 years old. Among children aged <5 years, the incidence of CHIKV-associated neurological disease was 77 per 100,000 person-years, compared with 20 per 100,000 for cerebral malaria and 7 per 100,000 for bacterial meningitis during the study period.

**Interpretation:** Although not previously recognized, CHIKV-associated neurological disease is common in coastal Kenya and a significant public health burden.

## INTRODUCTION

Chikungunya virus (CHIKV) is a positive sense RNA virus of the *Alphavirus* genus that was first discovered in Tanzania in 1953 [1]. CHIKV is transmitted between humans by *Aedes aegypti* and *Ae. albopictus* mosquitoes [2] which have facilitated its rapid global spread and the numerous chikungunya fever (CHIKF) epidemics reported to date [3]. In adults, CHIKF is characterized by abrupt onset of fever and debilitating muscle and joint pain, following an incubation period of about 2 to 10 days [4]. Most infections are self-limiting, but joint and musculoskeletal pain may persist for months to years in some individuals [4].

However, unlike adults, young children rarely present with musculoskeletal symptoms but are more likely to be hospitalised with CHIKV-associated neurological disease [5-10]. No specific therapeutics or licensed vaccines are available for CHIKF and, in the absence of pathognomonic clinical features, confirmatory diagnosis relies on laboratory detection of CHIKV in clinical samples [4].

CHIKV transmission is widely reported in Africa [3], with endemic febrile disease being particularly common in children [11-14]. In 2004, one of the largest CHIKF epidemics on record began in coastal Kenya and spread rapidly along the East African coast and to islands in the Indian Ocean [15-17]. During that epidemic, severe neurological manifestations of CHIKF were described in children in La Reunion [6, 18], including disease in neonates suggesting mother-to-child virus transmission [19, 20]. CHIKV-associated neurological disease has since been reported in other geographical settings [5, 8], except Africa.

Recent analysis identified a high, previously unrecognized, burden of endemic CHIKF in children presenting at outpatient primary care facilities in coastal Kenya [14]. Here, we used an established ward surveillance at Kilifi County Hospital on the north coast of Kenya, to estimate the incidence of CHIKV-associated neurological disease among children in this setting. Cerebrospinal fluid (CSF) samples taken for clinical reasons at the hospital are routinely stored and clinical records linked to the Kilifi Health and Demographic Surveillance System (KHDSS), allowing estimation of population incidence rates [21]. We therefore systematically tested these stored CSF samples to establish whether evidence of infection by CHIKV is common among children admitted to hospital with neurological illness in coastal Kenya.

## METHODS

### Clinical surveillance

Kilifi County Hospital is a referral public hospital in rural coastal Kenya with approximately 4,000 paediatric admissions annually. The hospital’ s catchment area includes the approximately 290,000 KHDSS residents who account for ∼40-50% of all admissions and are enumerated during household census rounds conducted every four months [21]. Malaria transmission is endemic in the demographic surveillance area with seasonal rains occurring in April-June and October-December [22]. The paediatric service at Kilifi County Hospital includes two wards; a 70-bed general ward and a 15-bed high dependency unit (HDU) staffed by research clinicians and nurses. The HDU admits children with serious illness requiring more intensive monitoring and management but has no ventilation facilities or renal replacement therapy. Electronic case records are kept for all admissions including demographics, vital signs, clinical history and examination, as well as routine laboratory investigations such as malaria blood slides, and blood and CSF culture. Each patient is assigned a discharge diagnosis by the attending clinician following review of the notes and results. All data are linked to the KHDSS database in real time by means of unique person identifiers.

For this study, all children aged <16 years whose illness required sampling and analysis of CSF were eligible for inclusion. The decision to collect CSF was made by clinicians. The same senior clinicians oversaw care throughout the period of surveillance. Where acute coma was present, the medical team considered the risks of lumbar puncture versus clinical benefits of diagnostic information from a clinical management perspective without reference to research considerations. Where clinically appropriate, delayed lumbar punctures were conducted after a period of treatment. Neuroimaging was not available on site during the period of surveillance. After investigations done for immediate clinical care, an aliquot of CSF was stored at -80°C and used for CHIKV testing by RT-PCR as described below. Ethical approval was provided by the Kenya Medical Research Institute Scientific and Ethics Review Unit (SSC No. 3296) and written informed consent provided by parents or guardians of all study participants.

### Detection of CHIKV infection

Total RNA was isolated from 100µl of each CSF sample using TRIzol™ Reagent (ThermoFisher). A published primer-probe set targeting the CHIKV non-structural protein 1 (nsP1) region [23] was then used to detect CHIKV viral RNA using the Taqman® Fast Virus RT-PCR kit (ThermoFisher) on a 7500 Real-Time PCR System (Applied Biosystems) in a 10µl reaction volume comprising: 3µl of 4x Taqman® Fast Virus 1-step master mix, 5µl RNA, and primers (CHIKV 874, CHIKV 961) and probe (CHIKV 899) [23] at final concentrations of 800nM and 200nM, respectively. RT-PCR conditions were; reverse-transcription at 50°C for 5 min, RT inactivation/initial activation 95°C for 20 sec and 45 cycles of denaturation at 95°C for 3sec and annealing/extension at 60°C for 30 sec [14]. A positive result was defined as a cycle threshold (Ct) value of <40, with viral RNA from a cultured CHIKV isolate and RT-PCR mastermix without template used as positive and negative controls, respectively [14].

### Statistical analysis

All children hospitalised during the study period and whose illness required sampling and analysis of CSF were included in the analysis. Demographic and clinical features were compared between CHIKV positive and CHIKV negative admissions using Chi2 test for categorical variables and Mann-Whitney U test for continuous variables. Cerebral malaria was defined as admission with a *Plasmodium falciparum* parasite density >2,500/μl of blood and a Blantyre Coma Score (BCS) <3, while impaired consciousness was defined as BCS 3 or 4 [22, 24]. Acute bacterial meningitis was defined as either: i) a positive CSF bacterial culture or latex agglutination test, or ii) bacteraemia accompanied by a CSF-to-blood glucose ratio <0.1, or iii) bacteraemia accompanied by CSF white blood cell count ≥50□×□10^6^ cells/L [25, 26]. We required evidence of bacteria in CSF or in blood in the case definition of meningitis to maximise specificity, since there are no data on glucose or CSF cell counts for CHIKV infection in our setting. Incidence of CHIKV-associated neurological disease among KHDSS residents was calculated by dividing the number of CHIKV positive admissions by the total person-years of observation during the 5-year study period and expressed per 100,000 person-years observed. For comparison, incidence estimates for cerebral malaria and acute bacterial meningitis were also calculated. Incidence rate ratios were estimated using Poisson regression models and compared between socio-demographic variables. All analyses were carried out in STATA/IC version 15.1 (StataCorp College Station, Texas, USA). We also examined the geographical distribution of cases of CHIKV infection, cerebral malaria, meningitis and other admissions across the KHDSS area by mapping (using QGIS version 3.10) the geolocations of homes where the children lived at the time of admission. These data on residence are collected and routinely updated by the KHDSS.

## RESULTS

Between January 2014 and December 2018, 18341 children aged <16 years were admitted at Kilifi County Hospital, of whom 4332 (24%) had CSF collected for routine investigations (Figure 1). The most common clinical indications for CSF collection were coma, impaired consciousness and seizures which together accounted for 90% of all CSF collections (Figure S1). Similar proportions of children with these lumbar puncture indications had CSF collected across the 5-year study period, suggesting a consistent pattern of clinical practice throughout (Figure S2). After acute investigations were done for immediate clinical care, stored CSF was available for 3980 (92%) of the 4332 admissions and these were screened for CHIKV infection (Figure 1).

**Figure 1.**
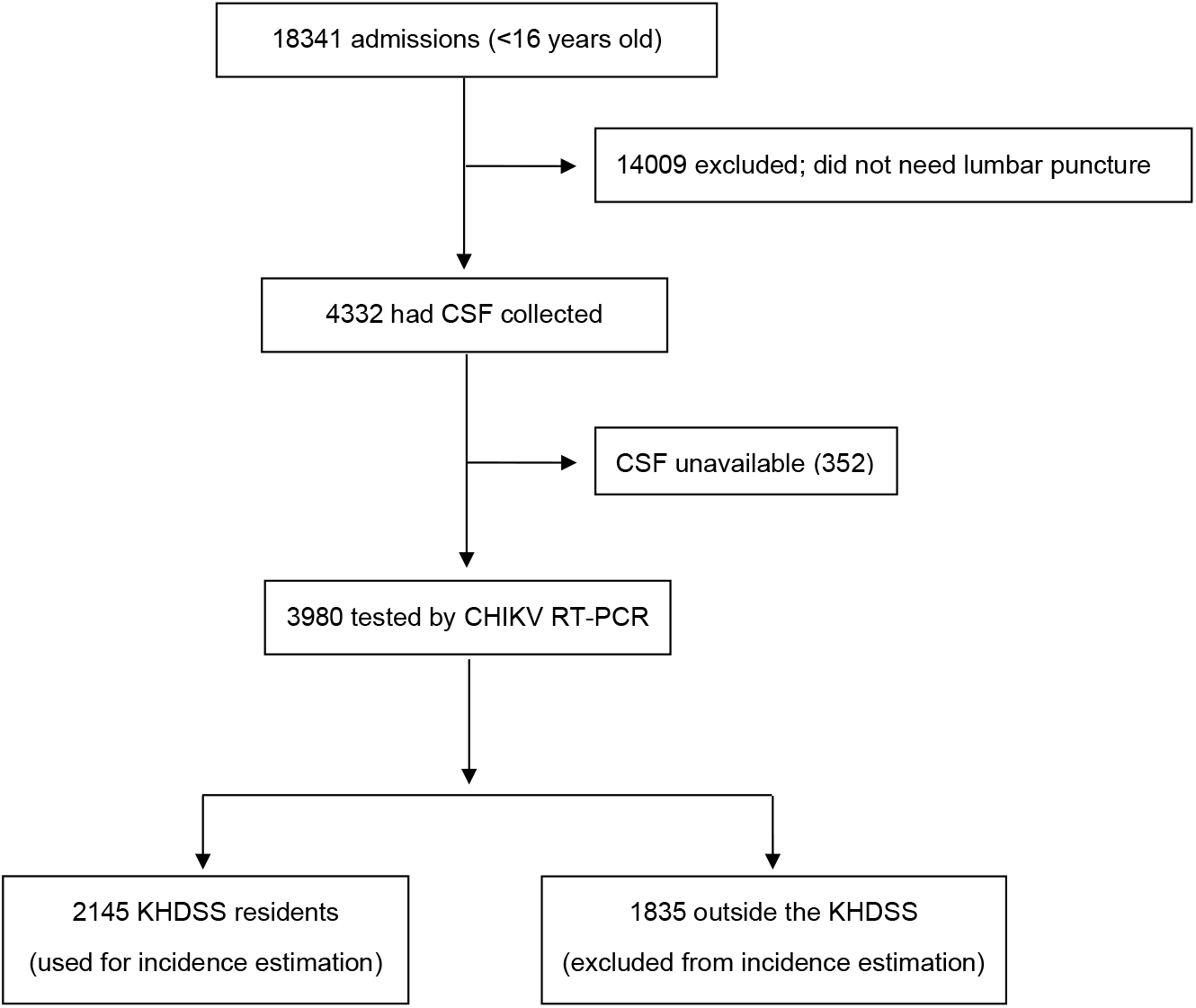

Of the 3980 admissions among children aged <16 years, 367 (9.2%, 95% CI 8.3, 10.2) were CHIKV RT-PCR positive. Most of these CHIKV infections (308 of the 367; 84%) were in children aged under 5 years (Table 1). RT-PCR assay cycle threshold values for CHIKV positive samples showed no correlation with age (Spearman’ s rho=-0.09, p=0.07). CHIKV RT-PCR positivity was highest in 2016 (18%), when an epidemic was reported in Kenya [27], and ranged between 4% to 9% in the other years (Table 1). CHIKV infection showed no association with HIV, bacteraemia or malaria parasitaemia at the time of admission (Table 1). Further, the distribution of clinical history and symptoms recorded at admission and laboratory investigations undertaken to inform clinical care was similar for CHIKV positive and CHIKV negative children (Table 1). Among children below 3 months of age, the majority (1352/1789; 75%) of CSF samples were taken from newborns in the first week of life in whom we observed high CHIKV positivity rates (8.7%, 95% CI 7.3, 10.3: Table S2) suggesting mother-to-child CHIKV transmission [28].

**Table 1:**
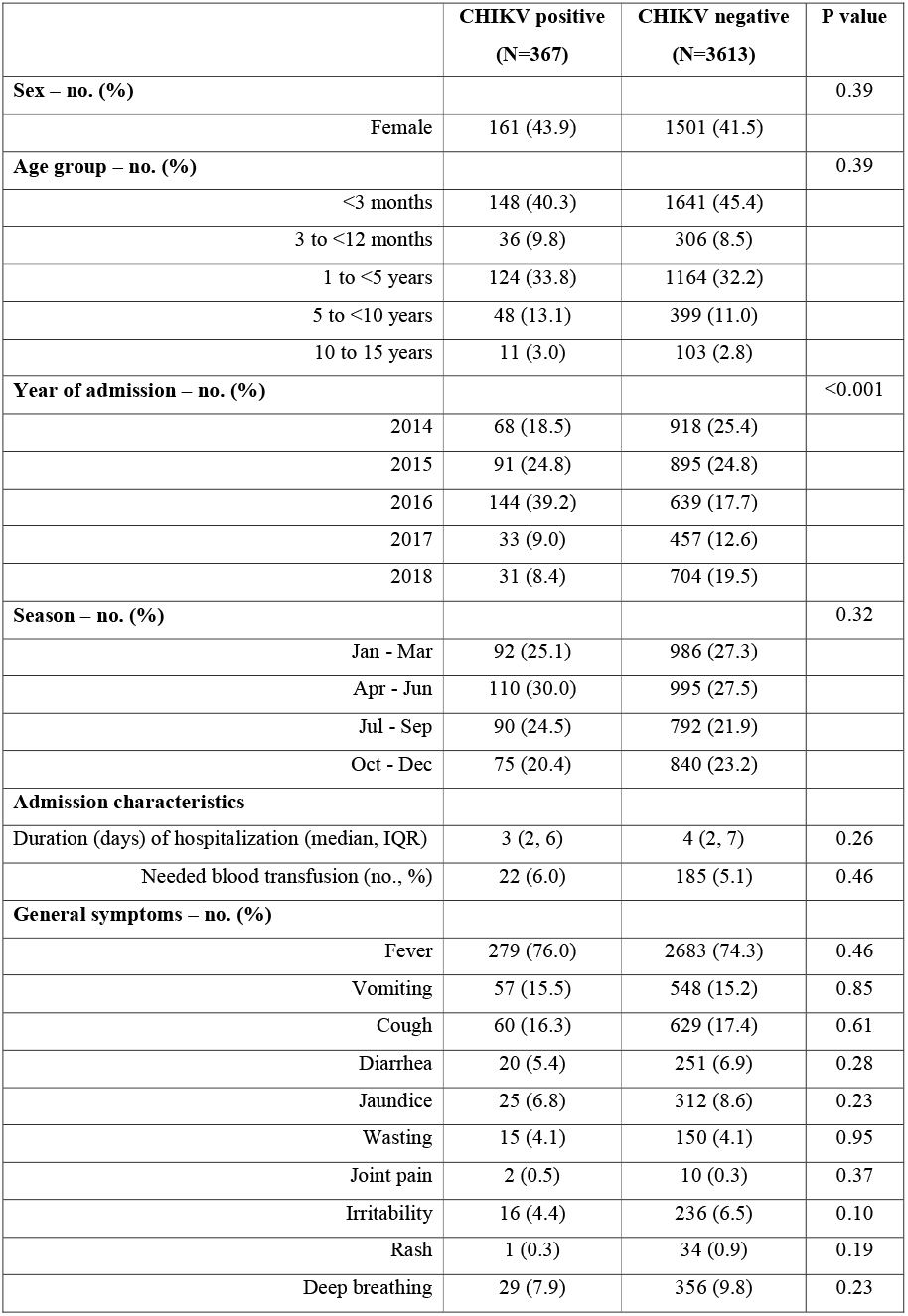
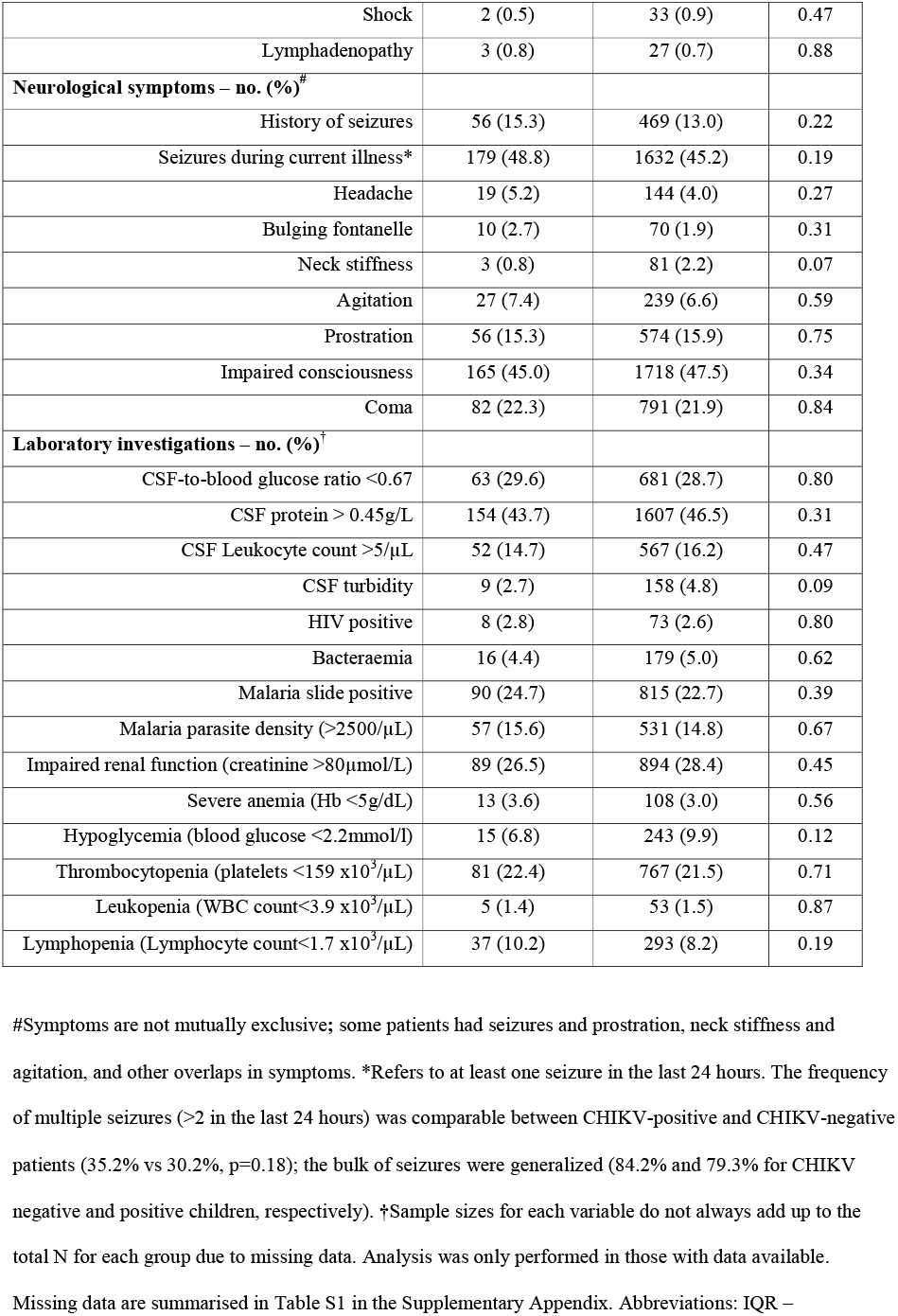
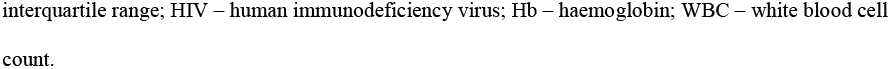
Demographic and clinical features of patients screened for CHIKV infection.

|  | **CHIKV positive**<br>**(N=367)** | **CHIKV negative**<br>**(N=3613)** | **P value** |
| --- | --- | --- | --- |
| **Sex – no. (%)** |  |  | 0.39 |
| Female | 161 (43.9) | 1501 (41.5) |  |
| **Age group – no. (%)** |  |  | 0.39 |
| <3 months | 148 (40.3) | 1641 (45.4) |  |
| 3 to <12 months | 36 (9.8) | 306 (8.5) |  |
| 1 to <5 years | 124 (33.8) | 1164 (32.2) |  |
| 5 to <10 years | 48 (13.1) | 399 (11.0) |  |
| 10 to 15 years | 11 (3.0) | 103 (2.8) |  |
| **Year of admission – no. (%)** |  |  | <0.001 |
| 2014 | 68 (18.5) | 918 (25.4) |  |
| 2015 | 91 (24.8) | 895 (24.8) |  |
| 2016 | 144 (39.2) | 639 (17.7) |  |
| 2017 | 33 (9.0) | 457 (12.6) |  |
| 2018 | 31 (8.4) | 704 (19.5) |  |
| **Season – no. (%)** |  |  | 0.32 |
| Jan - Mar | 92 (25.1) | 986 (27.3) |  |
| Apr - Jun | 110 (30.0) | 995 (27.5) |  |
| Jul - Sep | 90 (24.5) | 792 (21.9) |  |
| Oct - Dec | 75 (20.4) | 840 (23.2) |  |
| **Admission characteristics** |  |  |  |
| Duration (days) of hospitalization (median, IQR) | 3 (2, 6) | 4 (2, 7) | 0.26 |
| Needed blood transfusion (no., %) | 22 (6.0) | 185 (5.1) | 0.46 |
| **General symptoms – no. (%)** |  |  |  |
| Fever | 279 (76.0) | 2683 (74.3) | 0.46 |
| Vomiting | 57 (15.5) | 548 (15.2) | 0.85 |
| Cough | 60 (16.3) | 629 (17.4) | 0.61 |
| Diarrhea | 20 (5.4) | 251 (6.9) | 0.28 |
| Jaundice | 25 (6.8) | 312 (8.6) | 0.23 |
| Wasting | 15 (4.1) | 150 (4.1) | 0.95 |
| Joint pain | 2 (0.5) | 10 (0.3) | 0.37 |
| Irritability | 16 (4.4) | 236 (6.5) | 0.10 |
| Rash | 1 (0.3) | 34 (0.9) | 0.19 |
| Deep breathing | 29 (7.9) | 356 (9.8) | 0.23 |
| Shock | 2 (0.5) | 33 (0.9) | 0.47 |
| Lymphadenopathy | 3 (0.8) | 27 (0.7) | 0.88 |
| **Neurological symptoms – no. (%)<sup>#</sup>** |  |  |  |
| History of seizures | 56 (15.3) | 469 (13.0) | 0.22 |
| Seizures during current illness* | 179 (48.8) | 1632 (45.2) | 0.19 |
| Headache | 19 (5.2) | 144 (4.0) | 0.27 |
| Bulging fontanelle | 10 (2.7) | 70 (1.9) | 0.31 |
| Neck stiffness | 3 (0.8) | 81 (2.2) | 0.07 |
| Agitation | 27 (7.4) | 239 (6.6) | 0.59 |
| Prostration | 56 (15.3) | 574 (15.9) | 0.75 |
| Impaired consciousness | 165 (45.0) | 1718 (47.5) | 0.34 |
| Coma | 82 (22.3) | 791 (21.9) | 0.84 |
| **Laboratory investigations – no. (%)<sup>†</sup>** |  |  |  |
| CSF-to-blood glucose ratio <0.67 | 63 (29.6) | 681 (28.7) | 0.80 |
| CSF protein > 0.45g/L | 154 (43.7) | 1607 (46.5) | 0.31 |
| CSF Leukocyte count >5/μL | 52 (14.7) | 567 (16.2) | 0.47 |
| CSF turbidity | 9 (2.7) | 158 (4.8) | 0.09 |
| HIV positive | 8 (2.8) | 73 (2.6) | 0.80 |
| Bacteraemia | 16 (4.4) | 179 (5.0) | 0.62 |
| Malaria slide positive | 90 (24.7) | 815 (22.7) | 0.39 |
| Malaria parasite density (>2500/μL) | 57 (15.6) | 531 (14.8) | 0.67 |
| Impaired renal function (creatinine >80μmol/L) | 89 (26.5) | 894 (28.4) | 0.45 |
| Severe anemia (Hb <5g/dL) | 13 (3.6) | 108 (3.0) | 0.56 |
| Hypoglycemia (blood glucose <2.2mmol/l) | 15 (6.8) | 243 (9.9) | 0.12 |
| Thrombocytopenia (platelets <159 x10<sup>3</sup>/μL) | 81 (22.4) | 767 (21.5) | 0.71 |
| Leukopenia (WBC count<3.9 x10<sup>3</sup>/μL) | 5 (1.4) | 53 (1.5) | 0.87 |
| Lymphopenia (Lymphocyte count<1.7 x10<sup>3</sup>/μL) | 37 (10.2) | 293 (8.2) | 0.19 |

<sup>#</sup>Symptoms are not mutually exclusive; some patients had seizures and prostration, neck stiffness and agitation, and other overlaps in symptoms. \*Refers to at least one seizure in the last 24 hours. The frequency of multiple seizures (>2 in the last 24 hours) was comparable between CHIKV-positive and CHIKV-negative patients (35.2% vs 30.2%, p=0.18); the bulk of seizures were generalized (84.2% and 79.3% for CHIKV negative and positive children, respectively). <sup>†</sup>Sample sizes for each variable do not always add up to the total N for each group due to missing data. Analysis was only performed in those with data available. Missing data are summarised in Table S1 in the Supplementary Appendix. Abbreviations: IQR –
interquartile range; HIV – human immunodeficiency virus; Hb – haemoglobin; WBC – white blood cell count.

Overall mortality among all 18341 admissions aged <16 years was 9.0% (95% CI 8.6, 9.4; 1653 deaths). Mortality among the 4332 admissions that had CSF collected during the study period (Figure 1) was 3.2% (95% CI 2.7, 3.8; 139 deaths), compared with 10.8% (95% CI 10.3, 11.3; 1514 deaths) in those where CSF was not collected (Figure S3). A similar pattern was observed among newborns in the first week of life, where overall mortality was 17.7% (95% CI 16.5, 18.8), whereas mortality in those whose CSF was collected was 2.5% (39 deaths) compared with 25.3% (773 deaths) in those where CSF was not collected.

Overall case fatality among CHIKV positive children was 1.4% (95% CI 0.4, 3.2; 5 deaths of 367 CHIKV positive children) and 3.2% (95% CI 2.6, 3.8; 115 deaths) among CHIKV negative children (Table 1).

To estimate the incidence of CHIKV-associated neurological disease we took the numerator as CHIKV RT-PCR positive admissions among KHDSS residents (i.e., 207 cases were from KHDSS residents out of the 367 cases presenting to the hospital; Figure 2) and the denominator as the total person-years observed in the KHDSS during the 5-year study period for children aged <16 years (691,588 person-years; Table 2). The overall incidence of CHIKV-associated neurological disease within the KHDSS was 30 per 100,000 person-years (95% CI 26.1, 34.3) and showed no significant variation by sex or season (Table 2). Disease incidence was highest during the 2016 epidemic but a substantial number of presentations with CHIKV infection were also detected in other years (Table 2). CHIKV-associated neurological disease cases were distributed throughout the KHDSS area (Figure 2). A strong inverse relationship was observed between the incidence of CHIKV infection and age, estimated at 77 per 100,000 person-years in all children aged <5 years and 7 per 100,000 person-years among children aged ≥5 years (Table 2). During the same period, we calculated the corresponding incidences of cerebral malaria and bacterial meningitis in children aged <5 years to be 20 per 100,000 and 7 per 100,000 person-years, respectively (Table 2). The corresponding incidence of cerebral malaria and bacterial meningitis in children aged ≥5 years was 5 per 100,000 and 7 per 100,000 person-years, respectively (Table 2).

**Figure 2.**
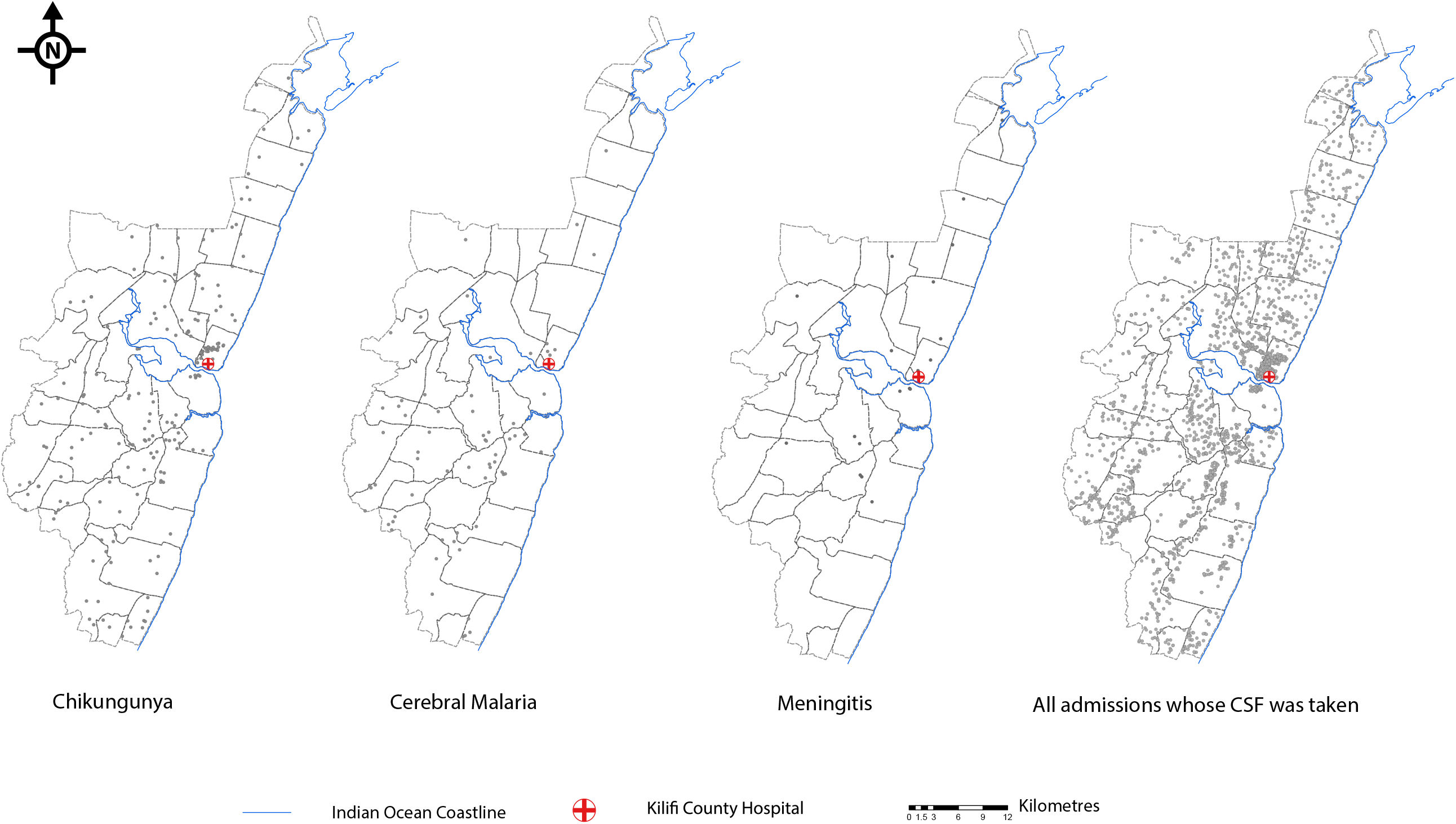

**Table 2:** Incidence of CHIKV-associated neurological disease among children aged <16 years within the KHDSS during the study period (2014-2018)

| Variable | Categories | Chikungunya (N=207) |  |  | Cerebral Malaria (N=68) |  |  | Acute Bacterial Meningitis (N=22) |  |  |
| --- | --- | --- | --- | --- | --- | --- | --- | --- | --- | --- |
|  |  | n / PYO | Incidence/100,000<br>(95% CI) | IRR<br>(95% CI) | n / PYO | Incidence/100,000<br>(95% CI) | IRR<br>(95% CI) | n / PYO | Incidence/100,000<br>(95% CI) | IRR<br>(95% CI) |
| Sex | Female | 95/341753 | 27.8 (22.7-34) | 1 | 33/341899 | 9.7 (6.9-13.6) | 1 | 7/341753 | 2 (1-4.3) | 1 |
|  | Male | 112/349835 | 32 (26.6-38.5) | 1.2 (0.6-2.4) | 35/350019 | 10 (7.2-13.9) | 1 (0.6-1.7) | 15/349835 | 4.3 (2.6-7.1) | 2.1 (0.9-5.1) |
| Age | <3 months | 77/10837 | 710.5 (568.3-888.4) | 1 | 0/10853 | - | - | 12/10851 | 110.6 (62.8-194.7) | 1 |
|  | 3 to <12 months | 19/33700 | 56.4 (36-88.4) | 0.1 (0-0.1) | 2/33755 | 5.9 (1.5-23.7) | 1 | 2/33750 | 5.9 (1.5-23.7) | 0.1 (0-0.2) |
|  | 1 to <5 years | 77/179432 | 42.9 (34.3-53.7) | 0.1 (0-0.1) | 42/179650 | 23.4 (17.3-31.6) | 3.9 (1-16.3) | 1/179697 | 0.6 (0.1-4) | 0 (0-0) |
|  | 5 to <10 years | 28/227125 | 12.3 (8.5-17.9) | 0 (0-0) | 20/227148 | 8.8 (5.7-13.6) | 1.5 (0.3-6.4) | 3/227227 | 1.3 (0.4-4.1) | 0 (0-0) |
|  | 10 to 15 years | 6/240495 | 2.5 (1.1-5.6) | 0 (0-0) | 4/240512 | 1.7 (0.6-4.4) | 0.3 (0.1-1.5) | 4/240519 | 1.7 (0.6-4.4) | 0 (0-0) |
| Year | 2014 | 41/138881 | 29.52 (21.7-40.1) | 1 | 15/138891 | 10.8 (6.5-17.9) | 1 | 4/138897 | 2.9 (1.1-7.7) | 1 |
|  | 2015 | 50/138008 | 36.2 (27.5-47.8) | 1.2 (0.8-1.9) | 24/138042 | 17.4 (11.7-25.9) | 1.6 (0.8-3.1) | 6/138062 | 4.3 (2-9.7) | 1.5 (0.4-5.3) |
|  | 2016 | 79/136770 | 57.8 (46.3-72) | 2 (1.3-2.9) | 12/136847 | 8.8 (5-15.4) | 0.8 (0.4-1.7) | 5/136877 | 3.7 (1.5-8.8) | 1.3 (0.3-4.7) |
|  | 2017 | 18/138861 | 13 (8.2-20.6) | 0.4 (0.3-0.8) | 7/138965 | 5 (2.4-10.6) | 0.5 (0.2-1.1) | 0/138998 | - | - |
|  | 2018 | 19/139067 | 13.7 (8.7-21.4) | 0.5 (0.3-0.8) | 10/139172 | 7.2 (3.9-13.4) | 0.7 (0.3-1.5) | 7/139209 | 5 (2.4-10.5) | 1.7 (0.5-6) |
| Season | Jan - Mar | 56/172895 | 32.4 (24.9-42.1) | 1 | 18/172967 | 10.4 (6.6-16.5) | 1 | 5/172996 | 2.9 (1.2-6.9) | 1 |
|  | Apr - Jun | 54/173137 | 31.2 (23.9-40.7) | 1 (0.7-1.4) | 15/173216 | 8.7 (5.2-14.4) | 0.8 (0.4-1.7) | 9/173245 | 5.2 (2.7-10) | 1.8 (0.6-5.4) |
|  | Jul - Sep | 53/172957 | 30.6 (23.4-40.1) | 0.9 (0.6-1.4) | 18/173043 | 10.4 (6.6-16.5) | 1 (0.5-1.9) | 3/173076 | 1.7 (0.6-5.4) | 0.6 (0.1-2.5) |
|  | Oct - Dec | 44/172599 | 25.5 (19-34.3) | 0.8 (0.5-1.2) | 17/172692 | 9.8 (6.1-15.8) | 0.9 (0.5-1.8) | 5/172726 | 2.9 (1.2-7) | 1 (0.3-3.5) |

PYO: person-years observed; IRR: incidence rate ratio

## DISCUSSION

We show that CHIKV is endemic in coastal Kenya, with viral RNA detected in approximately 9% of all CSF samples in a county referral hospital. We also find that the risk of CHIKV infection is highest in infants with disease being rare in older children, consistent with acquisition of immunity [14]. We show that CHIKV infections are common among newborns in our setting, within the first week of life, suggesting mother-to-child virus transmission as has been observed by others [19, 28]. None of the clinical or laboratory features available in our dataset could reliably distinguish CHIKV-associated illness from other causes of neurological disease, including meningism, seizures or coma, though fontanelle bulging was common among CHIKV positive neonates. Hence in the absence of specific laboratory diagnostics the disease burden of CHIKV among children with neurological illness is hidden.

92% of children had meningism, coma, seizures or mild reductions in Blantyre Coma Score that did not meet the threshold for coma as the indication for CSF collection. The mortality among CHIKV positive children was low, despite coma in 22% and depressed levels of consciousness in 47%. The decision to collect CSF via lumbar puncture was based on clinical priorities rather than research criteria. Even in those without obvious indications for CSF collection, the illness must have appeared sufficiently significant to clinicians to justify hospital admission and lumbar puncture. No specific treatment is available for CHIKV infection.

Diagnosis would improve antibiotic stewardship and avoid treatment costs. However, the sickest patients are likely to die before CSF can be collected as clinicians are less likely to collect CSF in the first 24 hours when a patient presents in acute coma. During the 5-year period of monitoring 632 children with coma did not have CSF collected, of whom 248 (i.e., 39.2%) died, compared to 530 children with coma who did have CSF collected, 20 of whom died (i.e., 3.7%) (see Figure S3). Children with coma may not have CSF collected acutely if clinicians are concerned about raised intracranial pressure, and on clinical recovery clinicians or families may decide to forego lumbar puncture. Our data suggest that children in whom CSF is not collected are at high risk of death, and it is therefore possible that CHIKV is a cause of mortality among these children. Future studies using IgM serology against CHIKV will help address this.

When comparing CHIKV-positive and CHIKV-negative children, none of the clinical symptoms or laboratory tests showed differences that were substantial enough for diagnostic use in clinical practice. Diagnostic uncertainty in the absence of CHIKV RT-PCR testing (or other laboratory tests such as CHIKV IgM serology) may be clinically challenging. For instance, treating children for possible culture-negative bacterial meningitis is costly and contributes to antimicrobial resistance in a hospital setting. Neuroimaging is difficult to access in our setting and usually requires costs to be borne by parents. Capacity for definitive molecular or serological testing is therefore required for confirmatory diagnosis of a disease that appears to be a common cause of admission in young children in coastal Kenya.

We observed CHIKV infections during and outside an epidemic year suggesting endemic CHIKV transmission in coastal Kenya. With the linkage to demographic surveillance, we estimate an incidence of 77 CHIKV-positive admissions per 100,000 person-years among children aged <5 years. This incidence is higher than recent incidences for bacterial meningitis which we estimate at 7 per 100,000 person-years, or for invasive bacterial diseases such as 37 per 100,000 for invasive salmonellosis or 3 per 100,000 for invasive pneumococcal disease [25, 29], and almost four times the incidence of cerebral malaria (20 per 100,000) in the same age group [22]. Following the reductions in severe malaria due to falling malaria transmission [22], and reductions in bacterial meningitis after vaccination [25], CHIKV may now be one of the most common causes of hospitalization with neurological disease among children aged <5 years in coastal Kenya.

The study has some limitations. We defined cases based on PCR detection of viral RNA which likely underestimated the true burden of CHIKV due to the short duration of viral RNA detection in tissues during CHIKV infections [30]. Furthermore, we are likely to have underestimated more severe CHIKV infections since CSF is unlikely to be collected from the most severely unwell children. We did not screen for other viruses that have been associated with neurological disease in Africa (e.g., adenovirus, herpesvirus and others [31]), and this warrants future study. Further studies are also needed to determine the nature and timing of mother-to-child CHIKV transmission, including maternal CHIKV screening as done by others [20]. The study was limited to a single geographical location on the Kenyan coast and our estimate of disease incidence was based on the KHDSS population as denominator, which generally accounts for 40-50% of all admissions at Kilifi County Hospital. However, CHIKV infections showed little geographical heterogeneity within the County, supporting generalizability of these results.

CHIKV mosquito vectors are widely distributed on the East African coast and reporting of cases of CHIKV infections in Africa is widespread. Despite this, surveillance for CHIKV in CSF samples has not been undertaken systematically in Africa and should now be an urgent priority to describe this previously unidentified public health burden. Such work will provide the foundation for future research on preventive and therapeutic interventions.

## Data Availability

All data are available in the main text or the supplementary materials.

## ACKNOWLEDGEMENTS

This work was commissioned by the National Institute for Health Research (NIHR) Global Health Research programme (16/136/33) using UK aid from the UK Government. The views expressed in this publication are those of the authors and not necessarily those of the NIHR or the Department of Health and Social Care.

GMW is supported by an Oak foundation fellowship and a Wellcome Trust grant (grant number 203077_Z_16_Z). This manuscript was submitted for publication with permission from the Director of the Kenya Medical Research Institute. This research was funded in part by the Wellcome Trust grant number 203077_Z_16_Z. For the purpose of Open Access, the author has applied a CC-BY public copyright licence to any author accepted manuscript version arising from this submission.

## AUTHORS’ CONTRIBUTIONS

DKN: Investigation, Formal Analysis, Methodology, Writing - review & editing

MO: Formal Analysis, Visualisation, Methodology, Writing - review & editing

LM: Data Curation, Writing - review & editing

SMK: Formal Analysis, Writing - review & editing

BO: Formal Analysis, Writing - review & editing

DOO: Investigation, Methodology, Writing - review & editing

GG: Investigation, Validation, Writing - review & editing

BK: Data Curation

HKK: Investigation, Writing - review & editing

JNG: Investigation, Writing - review & editing

ZRL: Investigation, Writing - review & editing

AD: Visualisation, Writing - review & editing

SM: Investigation, Writing - review & editing

CNA: Investigation, Validation, Writing - review & editing

SMT: Supervision, Writing - review & editing

MMH: Project Administration, Writing - review & editing

CRN: Supervision, Writing - review & editing

PB: Conceptualisation, Formal Analysis, Writing - review & editing

GMW: Conceptualisation, Funding acquisition, Supervision, Investigation, Project Administration, Formal Analysis, Writing - original draft, Writing - review & editing

