## Supplemental material for "Incidence of chikungunya virus infections among Kenyan children with neurological disease: a prospective cohort study"

**SUPPLEMENTARY INFORMATION**

**Table of contents**

1. Table S1: Missing data
2. Table S2: Demographic and clinical features of patients aged <3 months screened for CHIKV infection
3. Figure S1: Distribution of clinical indications for CSF collection
4. Figure S2: Annual distribution of admissions at Kilifi County Hospital stratified by clinical indication for CSF collection.
5. Figure S3: Distribution of deaths by clinical indication for CSF collection

|  | **CHIKV positive**  **(N=367)** | **CHIKV negative**  **(N=3613)** |
| --- | --- | --- |
| **Laboratory investigations** | **Number missing** | **Number missing** |
| CSF-to-blood glucose ratio <0.67 | 154 | 1244 |
| CSF protein > 0.45g/L | 15 | 161 |
| CSF Leukocyte count >5/µL | 14 | 119 |
| CSF turbidity | 36 | 312 |
| HIV positive | 83 | 768 |
| Bacteraemia | 4 | 28 |
| Malaria slide positive | 2 | 20 |
| Malaria parasite density (>2500/µL) | 2 | 20 |
| Impaired renal function (creatinine >80µmol/L) | 31 | 470 |
| Severe anemia (Hb <5g/dL) | 5 | 50 |
| Hypoglycemia (blood glucose <2.2mmol/l) | 145 | 1166 |
| Thrombocytopenia (platelets <159 x10^3^/µL) | 5 | 50 |
| Leukopenia (WBC count<3.9 x10^3^/µL) | 5 | 48 |
| Lymphopenia (Lymphocyte count<1.7 x10^3^/µL) | 5 | 51 |

**Table S1: Missing data.** The number of children missing data within each clinical category is shown for the respective variables included in the analysis presented in Table 1.

|  | **CHIKV positive**  **(N=148)** | **CHIKV negative**  **(N=1641)** | **P value** |
| --- | --- | --- | --- |
| **Sex – no. (%)** |  |  | 0.84 |
| Female | 64 (43.2) | 696 (42.4) |  |
| **Age group – no. (%)** |  |  | 0.67 |
| Day of birth | 29 (19.6) | 305 (18.6) |  |
| 1 to 7 days | 88 (59.5) | 930 (56.7) |  |
| 8 to 14 days | 8 (5.4) | 134 (8.2) |  |
| 15 to 28 days | 9 (6.1) | 132 (8.0) |  |
| 29 to 90 days | 14 (9.5) | 140 (8.5) |  |
| **Year of admission – no. (%)** |  |  | <0.001 |
| 2014 | 27 (18.2) | 336 (20.5) |  |
| 2015 | 32 (21.6) | 387 (23.6) |  |
| 2016 | 64 (43.2) | 354 (21.6) |  |
| 2017 | 17 (11.5) | 248 (15.1) |  |
| 2018 | 8 (5.4) | 316 (19.3) |  |
| **Season – no. (%)** |  |  | 0.10 |
| Jan - Mar | 42 (28.4) | 489 (29.8) |  |
| Apr - Jun | 57 (38.5) | 479 (29.2) |  |
| Jul - Sep | 24 (16.2) | 318 (19.4) |  |
| Oct - Dec | 25 (16.9) | 355 (21.6) |  |
| **Perinatal history – no. (%)** |  |  |  |
| Premature birth | 15 (10.8) | 186 (12.0) | 0.68 |
| Perinatal admission | 53 (38.1) | 519 (33.4) | 0.26 |
| Unable to breastfeed | 45 (32.4) | 560 (36.1) | 0.38 |
| Cried at birth | 109 (78.4) | 1120 (72.1) | 0.11 |
| Perinatal jaundice | 11 (7.9) | 146 (9.4) | 0.56 |
| Low birth weight (<1500g) | 5 (3.6) | 66 (4.2) | 0.71 |
| **Admission characteristics** |  |  |  |
| Duration (days) of hospitalization (median, IQR) | 4 (2, 7) | 4 (3, 8) | 0.39 |
| Needed blood transfusion (no., %) | 5 (3.4) | 68 (4.2) | 0.67 |
| **General symptoms – no. (%)** |  |  |  |
| Fever | 93 (62.8) | 984 (60.0) | 0.49 |
| Vomiting | 6 (4.0) | 69 (4.2) | 0.93 |
| Diarrhea | 1 (0.7) | 17 (1.0) | 0.67 |
| Jaundice | 19 (12.8) | 274 (16.7) | 0.22 |
| Irritability | 12 (8.1) | 168 (10.2) | 0.41 |
| Rash | 1 (0.7) | 14 (0.8) | 0.82 |
| Lymphadenopathy | 0 (0) | 1 (0.1) | 0.76 |
| **Neurological symptoms – no. (%)** |  |  |  |
| Seizures during current illness | 22 (14.9) | 211 (12.9) | 0.19 |
| Bulging fontanelle | 7 (4.7) | 30 (1.8) | 0.02 |
| Coma | 31 (20.9) | 320 (19.5) | 0.67 |
| **Laboratory investigations – no. (%)** |  |  |  |
| CSF-to-blood glucose ratio <0.67 | 28 (35.4) | 265 (26.4) | 0.08 |
| CSF protein > 0.45g/L | 119 (83.8) | 1266 (82.4) | 0.67 |
| CSF Leukocyte count >5/µL | 36 (25.3) | 349 (22.2) | 0.38 |
| CSF turbidity | 7 (5.5) | 100 (7.1) | 0.50 |
| HIV positive | 4 (3.3) | 32 (2.4) | 0.52 |
| Bacteraemia | 8 (5.4) | 102 (6.3) | 0.68 |
| Malaria slide positive | 0 (0) | 4 (0.2) | 0.55 |
| Hypoglycemia (blood glucose <2.2mmol/l) | 11 (13.2) | 210 (19.9) | 0.14 |
| Thrombocytopenia (platelets <159 x10^3^/µL) | 16 (11.0) | 195 (12.1) | 0.70 |
| Leukopenia (WBC count<3.9 x10^3^/µL) | 1 (0.7) | 20 (1.2) | 0.56 |
| Lymphopenia (Lymphocyte count<1.7 x10^3^/µL) | 2 (1.4) | 32 (2.0) | 0.61 |

**Table S2: Demographic and clinical features of patients aged <3 months screened for CHIKV infection**

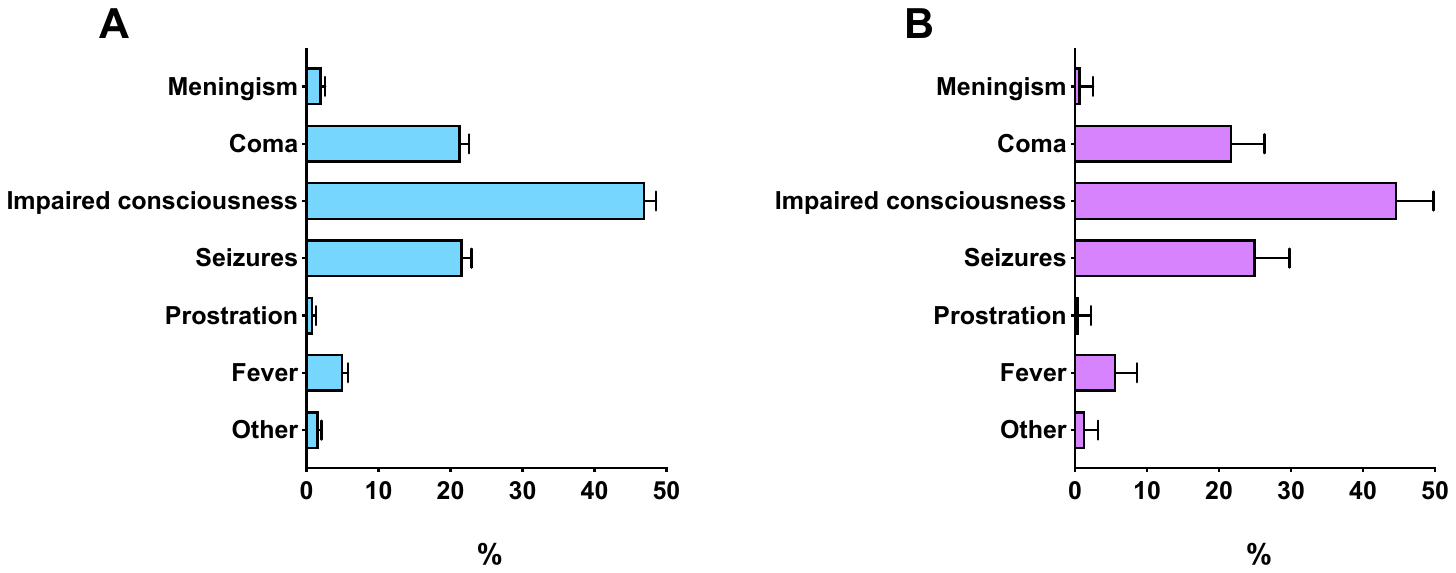

**Figure S1: Distribution of clinical indications for CSF collection.** The distribution of clinical indications for lumbar puncture among all 4332 admissions that had CSF collected during the study period are shown in panel A. In panel B, the distribution of clinical indications for the 367 children whose CSF were CHIKV positive is shown for comparison. The data are shown as percentages of the respective denominator (4332 in A, and 367 in B). The indications are organized according to a hierarchy, where we report the strongest indication for CSF collection taking the order of importance from top to bottom as: meningism; coma (BCS<3); impaired consciousness (BCS 3 or 4); seizures; prostration; fever; and other causes.

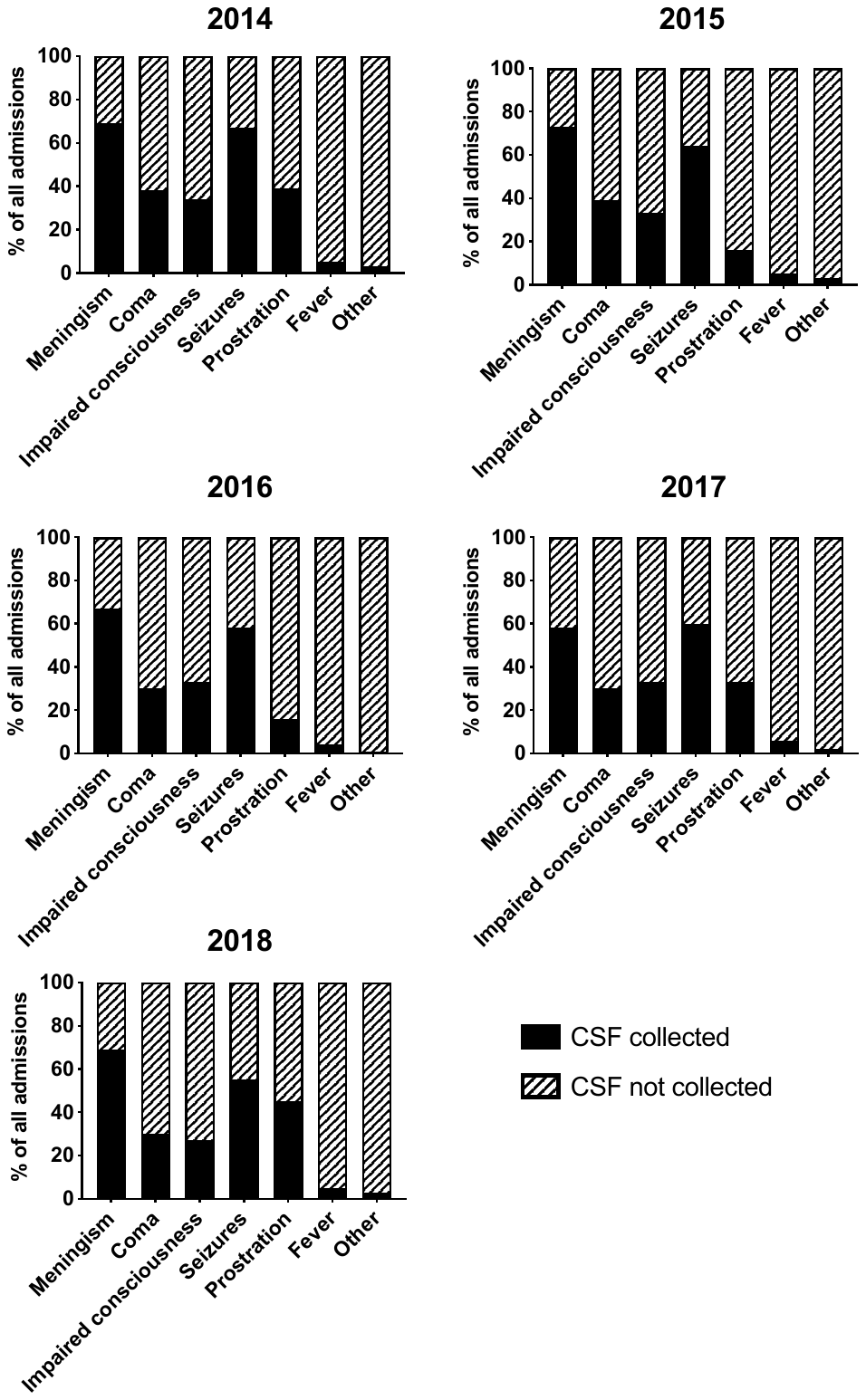

**Figure S2: Annual distribution of admissions at Kilifi County Hospital stratified by clinical indication for CSF collection.** The distribution of all 18341 children aged <16 years admitted at Kilifi County Hospital during the study period is shown, stratified by whether CSF was collected or not. The stacked bars for each clinical indication show the proportions whose CSF was collected or not collected and add up to 100% in each instance. The total number of admissions with each clinical indication over the 5-year study duration was: meningism (n=136), coma (n=2780), impaired consciousness (n=6428), seizures (n=1524), prostration (n=131), fever (n=4292) and others (n=3050).

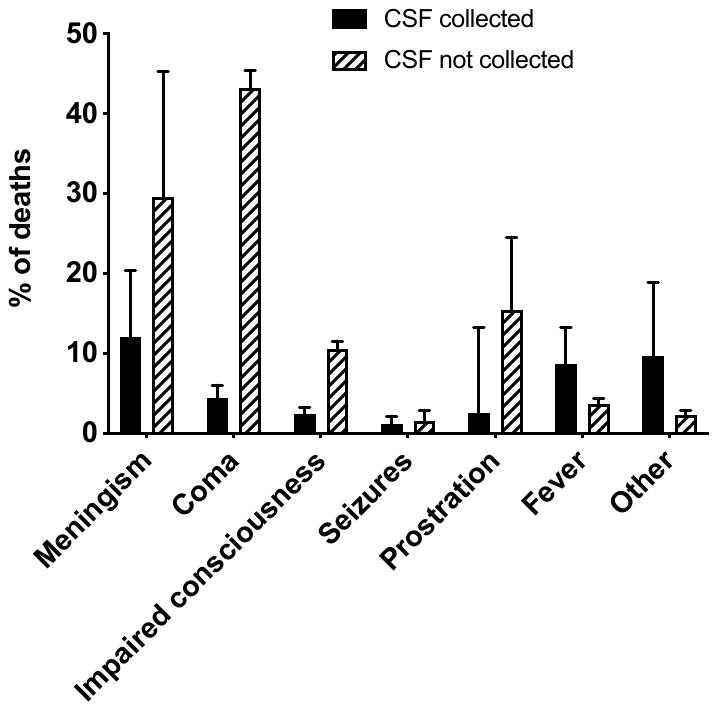

**Figure S3: Distribution of deaths by clinical indication for CSF collection.** There were 1653 (9.0%) deaths among all 18341 children aged <16 years admitted at Kilifi County Hospital during the study period. All deaths among children within each clinical indication are stratified by whether or not CSF was collected. The total number of deaths within each clinical indication are: meningism (n=24), coma (n=840), impaired consciousness (n=510), seizures (n=20), prostration (n=15), fever (n=169) and others (n=75).
